# An outbreak of SARS-CoV-2 in a public-facing office in England, 2021

**DOI:** 10.1101/2022.01.31.22269194

**Authors:** Barry Atkinson, Karin van Veldhoven, Ian Nicholls, Matthew Coldwell, Adam Clarke, Gillian Frost, Christina J. Atchison, Amber I. Raja, Allan M Bennett, Derek Morgan, Neil Pearce, Tony Fletcher, Elizabeth B Brickley, Yiqun Chen

## Abstract

Between August-September 2021, an outbreak of SARS-CoV-2, with an attack rate of 55% (22/40 workers), occurred in a public-facing office in England. To identify workplace and worker-related risk factors, a comprehensive investigation involving surface sampling, environmental assessment, molecular and serological testing, and worker questionnaires was performed in September – October 2021. The results affirm the utility of surface sampling to identify SARS-CoV-2 control deficiencies and the importance of evolving, site-specific risk assessments with layered COVID-19 mitigation strategies.

## Outbreak Setting and Attack Rates

The outbreak site was a three-storey public sector facility comprising three large open-plan offices, low-occupancy offices, meeting rooms, a canteen/lunchroom, and a small security office. Within the lower tier local authority area, the 7-day average SARS-CoV-2 incidence rate increased from 34/100,000 in mid-June to around 300/100,000 from late July onwards, and ranged between 268 and 331 weekly cases per 100,000 population during the outbreak period (Figure 1). Seventy-eight percent of local residents (aged ≥12 years) had received two doses of COVID-19 vaccines prior to the outbreak.

**Figure 1:**
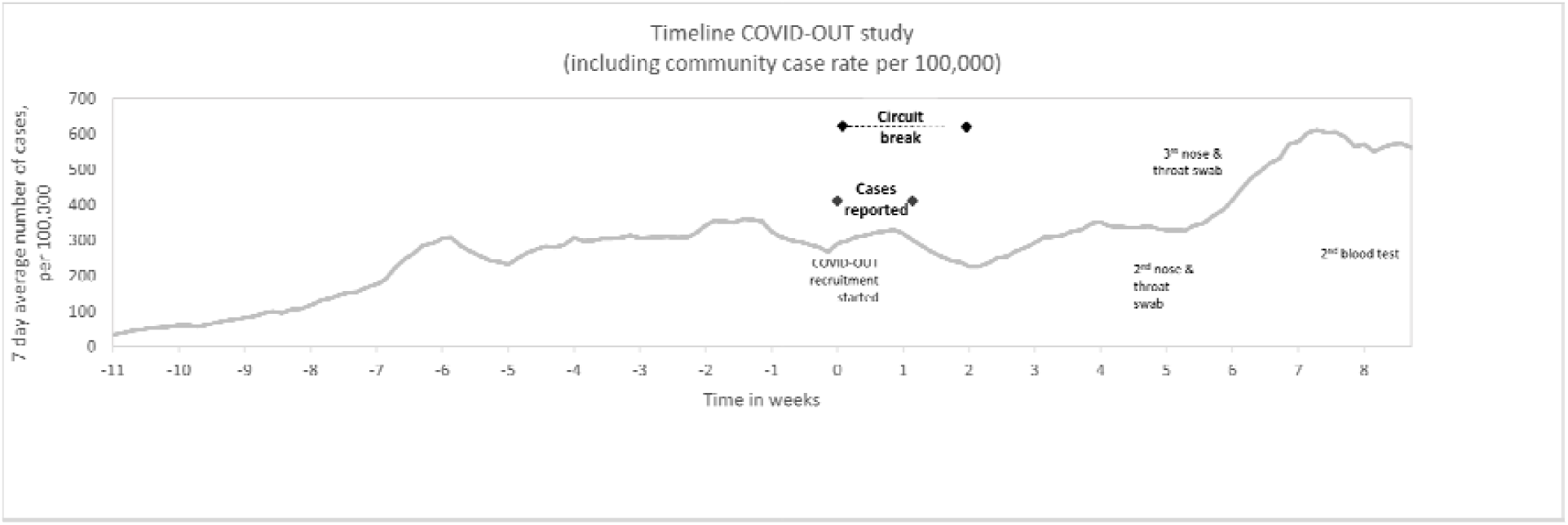
Timeline of the COVID-OUT investigation in a public-facing office, England, June to November 2021. Cases were identified between end of August and mid-September. Text/Arrows indicates key dates of the COVID-OUT study activities. The circuit-break, which was implemented at the moment of identification of the initial cluster of cases, lasted 2 weeks. The line chart represents the 7-day average case rate for the lower tier local authority area at which the company is based (publicly available from (https://coronavirus.data.gov.uk/); date of download 9 November 2021.

The workplace had a site specific COVID-19 prevention strategy in place based on a generic template provided by the parent organisation. Control measures included COVID-19 training for workers, single occupancy desks with 2m spacing, plastic dividers between workers and the public, commercial hand sanitizers, and a cleaning regimen including in between appointments with members of the public. Ventilation on site was predominately natural (i.e. manually opened windows) with some locally controlled air conditioning units and limited forced mechanical general ventilation.

The 40 workers included four (10.0%) cleaners, 30 (75.0%) office-based staff who had up to 15 face-to-face meetings with visitors per day, and six (15.0%) security staff who patrolled the facility on 30-minute rotating shifts, booked-in visitors, and operated shared equipment in a 7.8m^2^ office. The overall attack rate of SARS-CoV-2 was 55%, with 75% of cleaners (3/4), 47% of office staff (14/30), and 67% of security staff (4/6) testing positive. Between end of August and mid-September 2021, a ‘circuit break’ closure was implemented to stop transmission within the workplace.

### COVID-OUT Outbreak Investigation

Following notification, a comprehensive investigation by the COVID-19 Outbreak Investigation to Understand Transmission (COVID-OUT) study team, part of the PROTECT COVID-19 National Core Study on transmission and environment [1], was conducted in September and October 2021, using the previously described protocol [2].

Two rounds of surface sampling were performed using either Envirostik with Neutralising Buffer (Technical Service Consultants Ltd) or UTM® Viral Transport swabs (Copan) as appropriate. The first round of sampling was during the circuit break; the second round approximately two weeks later the site reopened. Quantitative reverse transcription polymerase chain reaction (qRT-PCR) for both ORF1ab and nucleocapsid (N) gene targets was performed in duplicate using the VIASURE SARS-CoV-2 Real Time PCR Detection Kit (CerTest) in accordance with the manufacturer’s instructions.

Out of 60 surfaces tested in the first round of sampling approximately 1-week after the first identified case, 10 (16.7%) were confirmed positives, and one (1.7%) was a suspected positive (Table 1). Five (8.3%) positive samples produced crossing threshold (Ct) values between 32.0-34.9; whole genome sequencing was attempted on these five samples, but only partial sequence was identified from a single sample (window handle). The data generated implied delta variant sequence but as <50% of the genome was recovered this could not be confirmed. The security office appeared to be a site of enhanced contamination, with six of nine (66.7%) samples from this location testing positive including three in the 32.0-34.9 Ct bracket. Based on these findings, enhanced cleaning was performed prior to the site re-opening, and routine cleaning procedures were updated to include window handle disinfection; maximum occupancy and cleaning regimens in the confined security office were also reassessed. Repeat surface sampling performed approximately 1-week after re-opening identified only one positive (2.4%) and one suspected positive (2.4%) sample, both near the assay’s limit of detection.

**Table 1:** Data collected from two rounds of surface sampling in the COVID-OUT investigation of an outbreak of SARS-CoV-2 in an office-based workplace, England, August to September 2021.

| Site sampling data |  |  |  |  |  |  |  |
| --- | --- | --- | --- | --- | --- | --- | --- |
|  |  | qRT-PCR result |  |  | qRT-PCR Mean Ct grouping |  |  |
| Time of sampling | Samples collected | Positive n (%) | Suspected n (%) | Negative n (%) | Ct <32.0 n (%) | Ct 32.0-34.9 n (%) | Ct >35.0<sup>a</sup> n (%) |
| Circuit-break | 60 | 10 (16.7%) | 1 (1.7%) | 49 (81.7%) | 0 (0.0%) | 5 (8.3%) | 55 (91.7%) |
| Reopening | 42 | 1 (2.4%) | 1 (2.4%) | 40 (95.2%) | 0 (0.0%) | 0 (0.0%) | 42 (100%) |
| Positive sample information |  |  |  |  |  |  |  |
| Location<br>Time of sampling | Description |  | Mean Ct Value<sup>b</sup> |  | Estimated copies per cm<sup>2c</sup> |  |  |
| Open plan office (2<sup>nd</sup> floor) Circuit-break | Window handle |  | 33.6 |  | 4,589.9 |  |  |
| Meeting room Circuit-break | Air vent |  | 37.6 (suspect<sup>d</sup>) |  | 25.1 |  |  |
| Meeting room Circuit-break | Desk + chair |  | 37.4 |  | 7.0 |  |  |
| Canteen Circuit-break | Table + chair |  | 34.2 |  | 35.2 |  |  |
| Security office Circuit-break | Radio (two-way) |  | 36.7 |  | 12.8 |  |  |
| Security office Circuit-break | Telephone |  | 34.6 |  | 583.1 |  |  |
| Security office Circuit-break | Key press |  | 34.5 |  | 148.1 |  |  |
| Security office Circuit-break | Control panel |  | 34.2 |  | 229.4 |  |  |
| Security office Circuit-break | Telephone |  | 35.1 |  | 214.2 |  |  |
| Security office Circuit-break | Desk + chair |  | 37.7 |  | 19.5 |  |  |
| Open plan office (ground floor) Circuit-break | Air conditioning unit |  | 37.6 |  | 13.1 |  |  |
| Canteen Reopening | Desk + chair |  | 37.6 (suspect<sup>d</sup>) |  | 1.1 |  |  |
| Canteen Reopening | Desk + chair |  | 37.2 |  | 1.7 |  |  |

<sup>a</sup>Includes samples with no SARS-CoV-2 RNA detected. <sup>b</sup>Mean Ct value for the N gene.
<sup>c</sup>Extrapolation from copies per reaction to copies per sample collected based on the dilution factor, then divided by recorded sampling area. <sup>d</sup>A 'suspected positive' is one where only a single replicate is positive in at least one target.

Ventilation was assessed using CO_2_ as a proxy; concentrations were determined by spot measurements when the facility re-opened, with continuous measurements logged over the subsequent two-week period in selected locations. These indicated that, although largely by natural means, ventilation conformed with current guidance [3], including in the security office where CO_2_ levels did not exceed 1200ppm and typically were <700ppm. At the time of the outbreak, the maximum occupancy of this office was two with security staff rotating positions regularly between this location and other areas within the building. Both occupancy levels for this office and security staff rotation patterns were modified upon reopening based on the findings of this study.

12 workers (100% office workers on regular day shift; 75% permanent contract) consented to participate in the COVID-OUT study, which included completing online questionnaires [4], two rounds of SARS-CoV-2 antibody testing, and three rounds of self-administered nose and throat swabs for qRT-PCR testing [2]. Of the 12 participants, five (41.7%) self-reported positive SARS-CoV-2 tests during the outbreak period; COVID-OUT serological testing confirmed all five were positive for both N- and S-specific antibodies against SARS-CoV-2 and two were positive by qRT-PCR. The non-cases were positive for S- but not N- antibodies, confirming that they had been vaccinated but not previously infected (Table 2). Prior to the outbreak, all participants, including the five cases, had received two doses of COVID-19 vaccines (date range of second dose: early May to mid-July in test-positives (cases) and mid-May to late July in non-cases; n=2 (18%) with Pfizer/BioNTech and n=9 (82%) with Oxford/AstraZeneca). All five cases were symptomatic for COVID-19 and presented with at least one of the following symptoms: fever, dry cough, productive cough, shortness of breath, and/or loss of taste or smell. Contact patterns were similar between cases and non-cases: roughly half reported >6 close contacts (i.e., during work, social, and essential activities) per day on average over the prior 14 days. Two-thirds of participants reported using face masks both at work and outside of work with a higher proportion of non-cases using face masks nearly all the time. Four participants (33%; two cases and two non-cases) reported direct physical or close contact with co-workers and members of the public. Of these, one worker (25%) reported a divider/screen between colleagues and three (75%) reported a divider/screen between themselves and members of the public. One participant reported no screen between themselves and colleagues or the public and reported that they could ‘sometimes’ maintain social distance with colleagues and ‘rarely’ with the public.

**Table 2.**
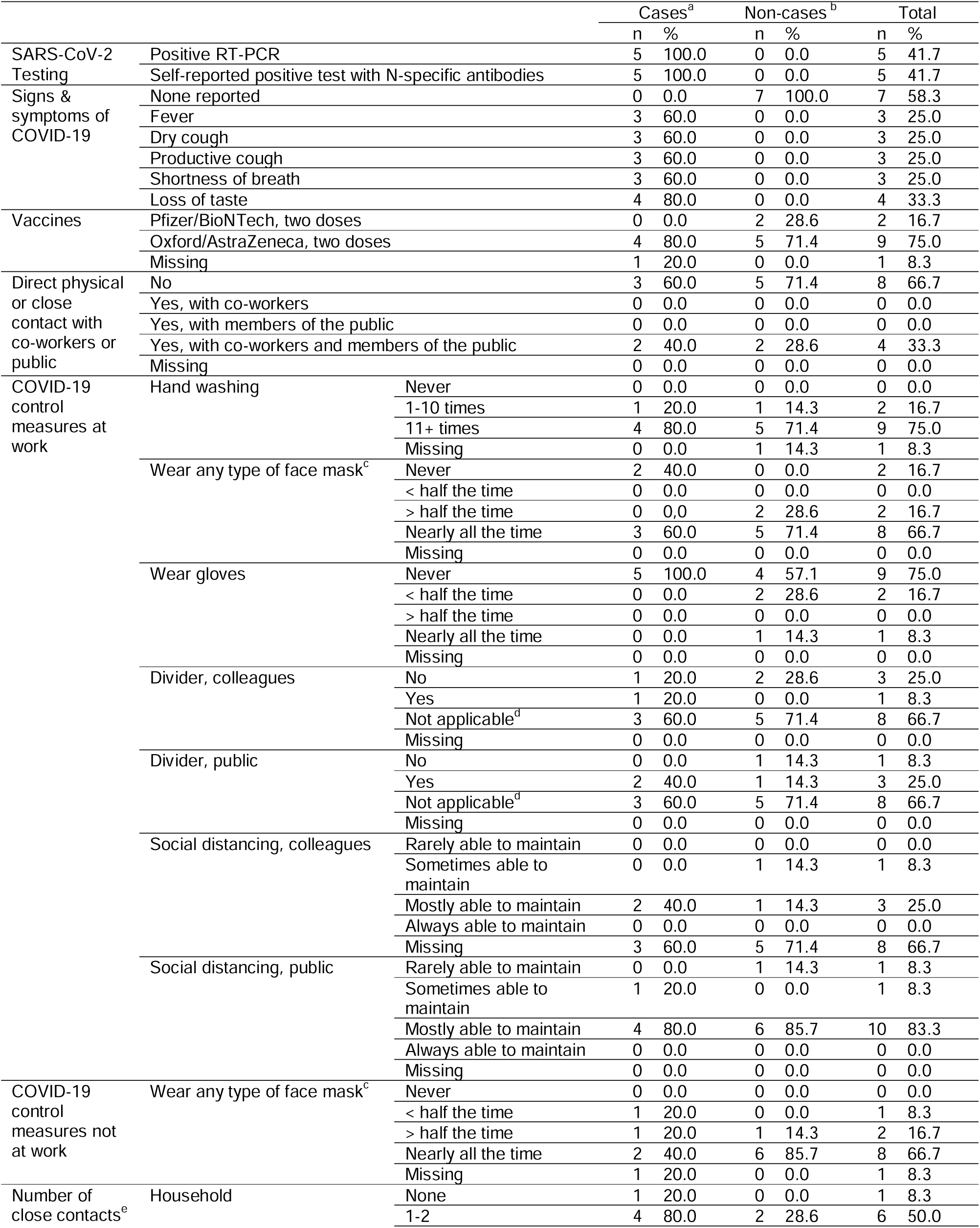
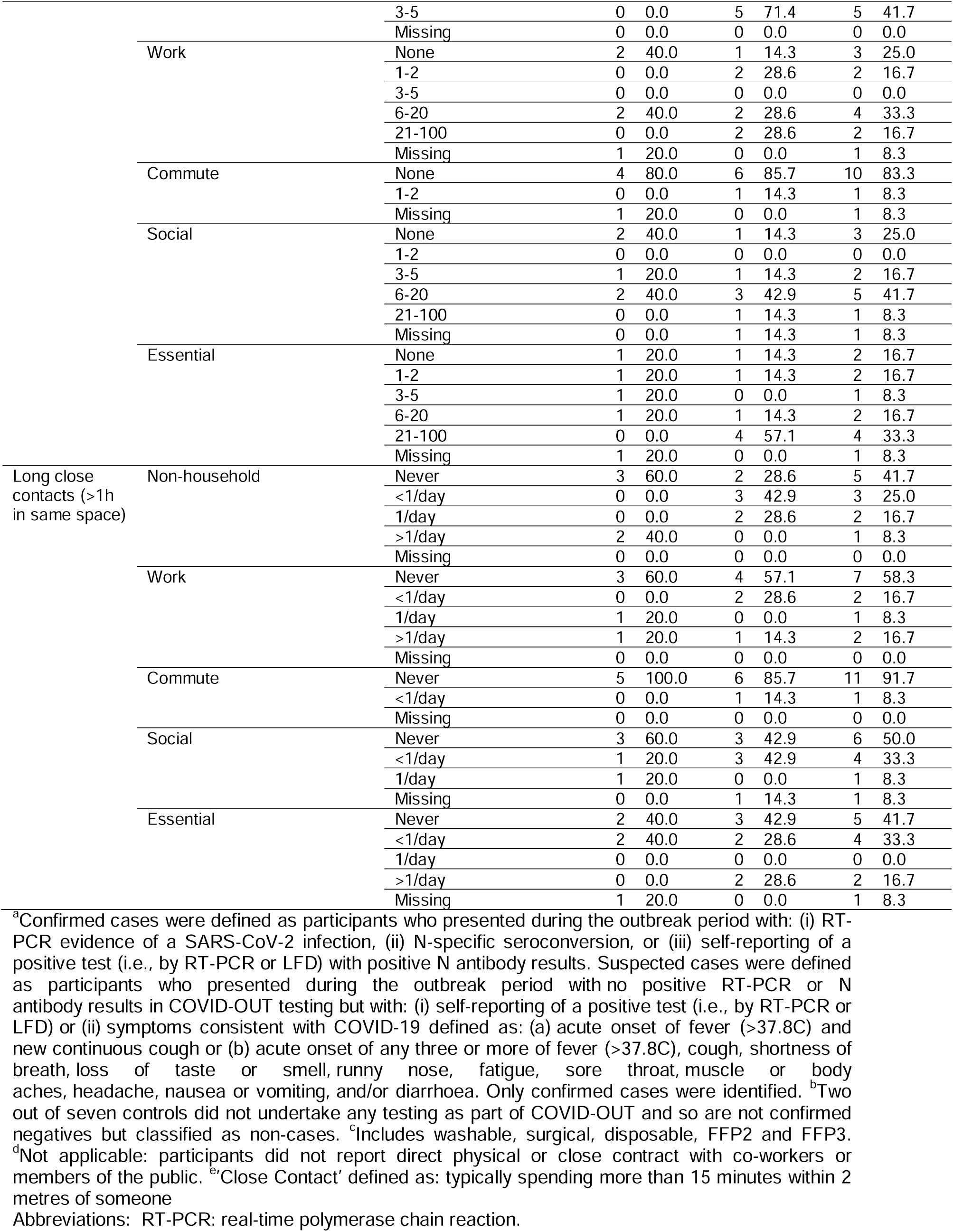
Test results, clinical presentation, and characteristics of participants in the COVID-OUT investigation of an outbreak of SARS-CoV-2 in an office-based workplace, England, August to September 2021.

## Discussion

In late August-September 2021, a public-facing office in England, with adherence to governmental COVID-19 control guidance and high vaccination coverage, experienced an outbreak of SARS-CoV-2 affecting 55% of the workforce. At the time of this outbreak, governmental guidance for workplaces in England prioritized policies for risk assessments, adequate ventilation, frequent cleaning, self-isolation, and communication/training; however, social distancing and face covering usage were no longer compulsory. Two weeks before the outbreak, government guidance was amended so fully vaccinated people did not need to self-isolate if they were identified as a close contact of someone with COVID-19.

Despite existing COVID-19 control measures, surface sampling identified potential deficiencies in routine disinfection procedures that informed targeted strengthening of infection control practices to support workplace re-opening. Similar to other SARS-CoV-2 outbreak studies, relatively low-level contamination was identified even in locations associated with recent occupancy of symptomatic people [5–10]. No positive surface sample yielded complete genome sequence suggesting degradation of the viral genome and a lack of transmission potential at the time of sampling. Similar investigations imply Ct values of <30 correlate with presence of infectious virus [11].

While vaccines remain highly effective for preventing severe COVID-19 illness and death, SARS-CoV-2 infections among fully vaccinated individuals in this outbreak are consistent with previous reports [12] and reinforce the importance of a layered SARS-CoV-2 transmission mitigation strategy prioritising ventilation and risk assessment-informed interventions, such as testing, social distancing, appropriate occupancy levels and transmission control measures (e.g. face masks), in addition to vaccination.

Overall, our findings highlight the need for evolving, site-specific risk assessments that adapt for changes in local community infection rates and recognize heterogeneity within a workplace in the risks associated with different workspaces (e.g. rooms with high occupancy) and worker roles (e.g. desk-based versus circulating staff). For public-facing workplaces, these assessments must inform suitable control measures to minimize potential close interactions of staff members with co-workers and visitors.

The limitations of this study warrant consideration. Surface sampling and participant testing performed closer to the peak of cases could yield a more representative indication of contamination within the facility and facilitate more informative genomic sequencing and epidemiological assessment [13]. Although workplace transmission appears likely, given the clustering of cases in some areas and positive environmental sampling, we were unable to clarify chains of transmission and determine whether cases may have been independently introduced from community sources. Notably, the worker participation rate in COVID-OUT was 30%, with an underrepresentation of male workers as well as cleaning and security staff. The small sample size and potential for selection bias limit our ability to evaluate individual risk factors within the workplace.

The ability to rapidly investigate SARS-CoV-2 outbreaks in workplaces and implement data-informed SARS-CoV-2 transmission mitigation measures is of importance for maintaining core societal functions during the COVID-19 pandemic. This is particularly relevant to workplaces with public-facing elements that have a dynamic population with an elevated risk of virus introduction and onward transmission. Mechanisms to encourage workplaces to report potential outbreaks as early as practicable and engage with research studies, like the one presented here, should be prioritized to further our understanding of transmission and to provide safer work environments.

## Copyright

© Crown copyright (2021), Health and Safety Executive. This is an open access article distributed under the terms of the Open Government Licence v3.0, which permits re-use, distribution, reproduction and adaptation, provided the original work is properly cited.

## Data Availability

All data produced in the present study are available upon reasonable request to the authors

## Acknowledgments

The authors wish to acknowledge Antony Spencer, Vince Sandys, Joan Cooke, Gary Dobbin, Hannah Higgins, Chris Keen, Alice Graham and Helen Collins for their efforts in recruitment and data collection supporting the COVID-OUT project, and for assisting with this investigation. The authors further acknowledge the assistance of the UKHSA Genomic Team for sequencing samples of interest from this study.

## Declarations

### Ethical approval

The COVID-OUT study has been approved by the NHS North East Research Ethics Committee (Reference 20/NE/0282).

### Data availability statement

The data that support the findings of this study are presented in this report. Additional data relating to the wider COVID-OUT project will be published and catalogued within the NCS PROTECT repository available at https://sites.manchester.ac.uk/covid19-national-project/.

## Authors’ contributions

**Barry Atkinson:** Conceptualisation, methodology, validation, formal analysis, investigation, data curation, writing – original draft, writing – review and editing, and supervision.

**Karin van Veldhoven:** Conceptualisation, methodology, validation, formal analysis, data curation, writing – original draft, writing – review and editing.

**Ian Nicholls:** Validation, formal analysis, investigation, data curation, writing – review and editing.

**Matthew Coldwell:** Conceptualisation, methodology, validation, formal analysis, investigation, data curation, writing – original draft, writing – review and editing, supervision.

**Adam Clarke:** Validation, formal analysis, investigation, data curation, writing – review and editing.

**Gillian Frost:** Validation, data curation, writing – original draft, writing – review and editing.

**Christina J. Atchison:** Methodology, writing – review and editing.

**Amber I. Raja:** Methodology, writing – original draft, writing – review and editing.

**Allan M Bennett:** Conceptualisation, methodology, validation, writing – original draft, writing – review and editing, supervision, project administration and funding acquisition.

**Derek Morgan:** Conceptualisation, methodology, writing – review and editing, supervision, project administration and funding acquisition.

**Neil Pearce:** Conceptualisation, methodology, writing – review and editing, project administration and funding acquisition.

**Tony Fletcher:** Conceptualisation, methodology, writing – review and editing.

**Elizabeth B Brickley:** Conceptualisation, methodology, writing – original draft, writing – review and editing, and supervision.

**Yiqun Chen:** Conceptualisation, methodology, validation, investigation, data curation writing – original draft, writing – review and editing, supervision, and project administration.

